# Applying mixture model methods to SARS-CoV-2 serosurvey data from Geneva

**DOI:** 10.1101/2021.07.19.21260410

**Authors:** Judith A Bouman, Sarah Kadelka, Silvia Stringhini, Francesco Pennacchio, Benjamin Meyer, Sabine Yerly, Laurent Kaiser, Idris Guessous, Andrew S Azman, Sebastian Bonhoeffer, Roland R Regoes

**Author notes:** Corresponding authors: Judith A. Bouman and Roland R. Regoes, (,).

## Abstract

Serosurveys are an important tool to estimate the true extent of the current SARS-CoV-2 pandemic. So far, most serosurvey data have been analysed with cut-off based methods, which dichotomize individual measurements into sero-positives or negatives based on a predefined cutoff. However, mixture model methods can gain additional information from the same serosurvey data. Such methods refrain from dichotomizing individual values and instead use the full distribution of the serological measurements from pre-pandemic and COVID-19 controls to estimate the cumulative incidence. This study presents an application of mixture model methods to SARS-CoV-2 serosurvey data from the SEROCoV-POP study from April and May 2020 in Geneva (2766 individuals). Besides estimating the total cumulative incidence in these data (8.1% (95% CI: 6.8% - 9.8%)), we applied extended mixture model methods to estimate an indirect indicator of disease severity, which is the fraction of cases with a distribution of antibody levels similar to hospitalised COVID-19 patients. This fraction is 51.2% (95% CI: 15.2% - 79.5%) across the full serosurvey, but differs between three age classes: 21.4% (95% CI: 0% - 59.6%) for individuals between 5 and 40 years old, 60.2% (95% CI: 21.5% - 100%) for individuals between 41 and 65 years old and 100% (95% CI: 20.1% - 100%) for individuals between 66 and 90 years old. Additionally, we find a mismatch between the inferred negative distribution of the serosurvey and the validation data of pre-pandemic controls. Overall, this study illustrates that mixture model methods can provide additional insights from serosurvey data.

## Introduction

Serological surveys (serosurveys) are an important tool to estimate the cumulative incidence of SARS-CoV-2 infections in various geographic locations or risk groups during the current pandemic [1]. Based on the estimated cumulative incidence, one can even calculate several related parameters such as: the ascertainment rate, i.e. the fraction of cases detected, the relative risk of infection for sub-groups [2], and the infection fatality rate [3]. In 2020, many serosurveys have been conducted in a wide variety of geographic locations [4]. The vast majority of these serosurvey studies have been analysed with cutoff-based methods, meaning that each individual serological measurement has been dichotomized into sero-negative or positive based on a predefined cutoff value. This cutoff value has been defined based on a receiver operating characteristic (ROC) curve constructed from samples from pre-pandemic controls and known SARS-CoV-2 infections.

The cutoff-based method for analysing serosurveys has two main challenges. Firstly, the cutoff depends on validation data from known SARS-CoV-2 infections, which are often not representative of the full spectrum of possible infections. Instead, cases used for the validation data are, especially at the beginning of an epidemic, biased towards severe infections and early convalescent periods [5]. However, it is known that disease severity influences the antibody level after infection [6] and that antibody levels wane over time [7]. This can lead to overly confident estimates of the sensitivity and specificity of the serological test and therefore bias the estimated cumulative incidence. Secondly, the amount of information obtained from the serosurvey is reduced by dichotomizing the continuous measurements. As a result, the cutoff-based method does not allow to differentiate between several types of SARS-CoV-2 infections (for instance mild and severe infections), nor to detect or correct for a possible mismatch between the cases included in the validation data and those in the serosurvey.

Both of the posed challenges can be circumvented by using mixture model methods. Instead of dichotomizing the individual serological observations, mixture model methods estimate the cumulative incidence directly based on the full distribution of serological measurements for the pre-pandemic controls and known SARS-CoV-2 infections [8]. As a result, this inference framework can also be used to determine whether the cases included as positive COVID-19 controls are a good representation of the cases in the serosurvey data or whether cases with a distinct distribution of serological measurements (such as measurements from individuals with an asymptomatic or mild infection) are missing in the validation data. Moreover, mixture model methods allow to use multiple distinct distributions of cases separately in the analysis. Even though mixture models have been successfully applied to serosurvey data for several pathogens [9]–[12], they are rarely used to analyse serosurvey data from SARS-CoV-2 studies [13].

In this study, we apply mixture model methods to serosurvey data from the SEROCoV-POP study that was performed in Geneva in April and May of 2020 [2]. In addition to corroborating previous estimates of the cumulative incidence for these data (4.6 % in first week (95% CI: 2.4%-8.0%) to 10.9 % in the fifth week (95% CI: 8.2%-13.9%)) – we estimated a cumulative incidence of 8.1% (95% CI: 6.8% - 9.8%) over the whole period of sampling –, our aim is to show how mixture model methods can be used to extract more information from serosurveys. We use an extended mixture model that takes into consideration the distribution of antibody levels of both hospitalized COVID-19 patients and outpatients. This results in an estimate of what we call the indirect indicator of severity, which is defined as the fraction of individuals in the serosurvey that display a distribution of antibody levels similar to that of the hospitalized patients in the control data. This fraction is not a direct estimate of the fraction of cases in the serosurvey that were treated in a hospital, as the validation data does not contain positive control data from asymptomatic and mild cases. Therefore, we rather refer to this quantity as the indirect indicator of disease severity.

## Methods

### Data

We used the pre-pandemic and COVID-19 control data from Meyer et al as the validation data in this study [14]. The pre-pandemic control data consists of 326 samples, 276 of these originated from adults and 50 from children [14]. They were collected in 2013, 2014 and 2018 at the University Hospitals of Geneva [14]. 84 of the samples came from healthy individuals and 242 from patients consulting the hospital [14]. The COVID-19 control data was collected from 181 individuals at the University Hospitals of Geneva. The severity of their infection is indicated by either ’hospitalized’ (n=91) or ’outpatient’ (n=90) [14]. Both hospitalized and outpatient individuals displayed at least mild symptoms.

We also used data from the SEROCoV-POP study from April and May 2020 from Stringhini et al [2]. Each week of the study, 1300 participants of the Bus Santé study were invited to participate via email and were asked to invite any household members [2]. The Bus Santé study is an annual cross-sectional study of adults residing in Geneva state (Switzerland) that, at the time of the study, had 17 225 participants on record [2], [15]–[18]. The invitation process resulted in the participation of 2766 individuals of which 52.6% are female [2]. Individuals aged between 50 and 64 were over-represented compared to the general population of Geneva and the age groups 5-9, 20-49 and 80-104 were under-represented [2]. Recruited participants have a higher educational level than the general population of Geneva [2]. The data from the 2766 recruited participants contains age, measured IgG OD ratio of the Euroimmun SARS-CoV-2 serological assay and sex. Additionally, the household structure between the individuals is indicated.

The serological assay measurements for all sera in both datasets were obtained with the Euroimmun SARS-CoV-2 serological assay which quantifies the IgG anti-bodies against the S1-domain of the spike protein of SARS-CoV-2 [14]. The IgG ratio is the result of the immunoreactivity of the sample measured at an optical density of 450 nm (OD450) divided by the OD450 of the calibrator [14], [19].

### Mixture model methods

We have assembled all observations of the SEROCoV-POP study from April and May and apply the mixture model described by Bouman et al [8]. All analyses are performed in R [20]. The basic mixture model maximizes the likelihood equation 1. Here, *U* is the vector of observed IgG OD ratios in the serosurvey data, *σ* is a binary vector of length *n* with their underlying true serological status (1 for past infection and 0 for no past infection). The probabilities *p*(*U*_*i*_|*σ*_*i*_ = 0) and *p*(*U*_*i*_|*σ*_*i*_ = 1) capture the empirical distributions of IgG OD ratios for the pre-pandemic and COVID-19 control measurements, and *π* is the cumulative incidence. The empirical distributions are obtained by smoothing the observed distributions. This is done with the ‘density’-function in R using the default kernel setting, ’gaussian’. [20]. The use of this smoothing function has been validated with simulated data in Bouman et al [8]. The specific smoothing algorithm does influence the results, even though the differences are small. For example, the kernel ‘cosine’ results in a point estimate of the cumulative incidence of 8.7 % (95%*CI* : 6.9% − 10.3%). More extensive data would allow us to determine the antibody distribution more reliable.

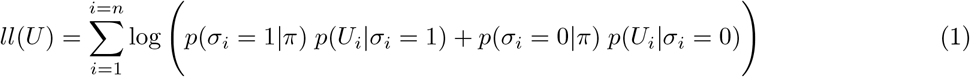

The likelihood is extended for the model where the outpatient and hospitalized cases are estimated separately, see equation 2. Here, *π*_*out*_ is the cumulative incidence of outpatient cases and *π*_*hosp*_ the cumulative incidence of hospitalized cases, *σ*_*i*_ can be 0 (no past infection), 1 (past outpatient infection) or 2 (past hospitalized infection).

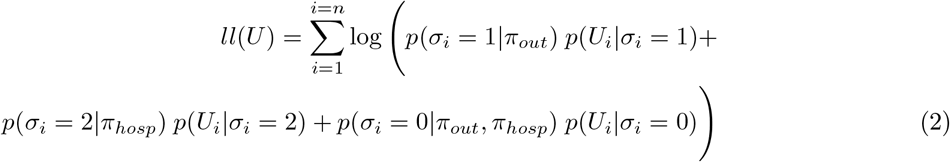

The 95% confidence intervals are estimated by bootstrapping the control distributions as well as the observations from the serosurvey. The various mixture models are compared with a likelihood ratio test.

We applied the extended model described above to the serosurvey data segregated into three age categories: 5-40 years, 41-65 years and 66-90 years. Even though the ages of the outpatient and hospitalized case distributions are significantly different, we used the whole distribution of both distributions for these analyses.

#### Testing for a mismatch between serosurvey and validation data

To test if there is a mismatch between the observed serosurvey data and the validation data, we extend Equation 2 with an additional class (see equation 3). Thus, *σ* can now take one of four categorical values where the new one represents an additional, yet unknown, category of cases. The distribution of this additional category (*p*(*U*_*i*_|*σ*_*i*_ = 3)) is modelled to be a normal distribution, where the mean and standard deviation are under optimization.

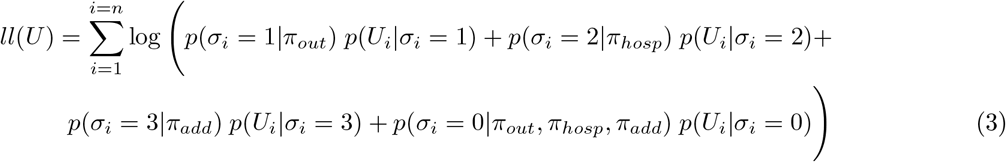

This model is then compared to the model of Equation 2 to test if the additional distribution has significantly improved the likelihood of observing the serosurvey data.

We have also used an adjusted version of the method described above, where we summarized all observations below 0.34 into a point mass for the empirical distribution. The value of 0.34 is two standard deviations larger than the mean of the inferred mismatch in the distribution of pre-pandemic controls, to make sure that this mismatch is not included in the new distributions. The model is then performed with these distributions instead of the original empirical distributions of the negative and positive controls.

## Results

### Distributions of IgG OD Ratios significantly differ for hospitalized and outpatient SARS-CoV-2 positive controls

Meyer et al. (2020) validated the diagnostic accuracy of the Euroimmun SARS-CoV-2 IgG and IgA immunoassay for SARS-CoV-2 infection [14]. For this validation, they used a pre-pandemic negative control group (Negative controls, 326 individuals) and two clinically distinguishable positive control groups: individuals who were hospitalized in the University Hospitals of Geneva (COVID-19 hospitalized, 91 individuals), and individuals who were treated in outpatient clinics (COVID-19 outpatients, 90 individuals). All positive controls tested positive for SARS-CoV-2 by PCR and showed at least mild symptoms. The observed IgG OD ratios of the Euroimmun SARS-CoV-2 immunoassay are shown in Figure 1 for the negative controls and both groups of positive controls. The distribution of the IgG OD ratios for the hospitalized positive controls is significantly different from the outpatient positive controls (two-sample Wilcoxon test, p-value = 1.122e-05).

**Figure 1:**
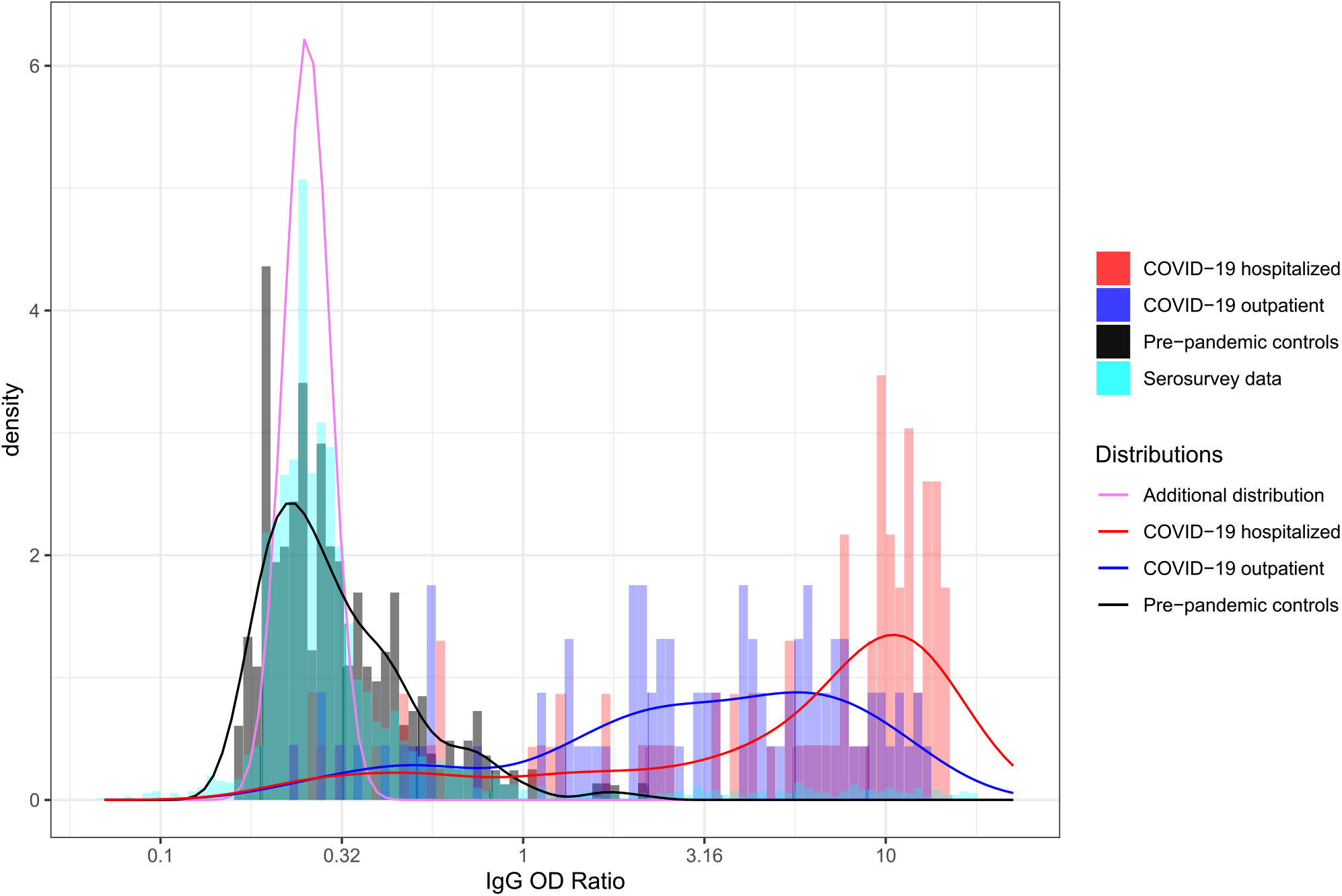
Histograms of IgG OD ratios of the Euroimmun SARS-CoV-2 IgG from the SEROCoV-POP study from April to May [2] and the validation data from Meyer et al [14]. Solid lines indicate the empirical distributions. The purple solid line shows the inferred additional distribution that is an indication of the mismatch between the pre-pandemic controls and the serosurvey data.

### Model that separately estimates the cumulative incidence for hospitalized and outpatient control data is significantly better than model based on one type of controls only

The significant difference between the distributions of the IgG OD ratios for the hospitalized and the outpatient controls allows the mixture model method to simultaneously estimate the cumulative incidence of both types of cases in the data from the SEROCoV-POP study from April and May 2020 (see Equation 2). We find a cumulative incidence of 4.0% (95% CI: 0.8% - 7.4%)) for cases with a distribution of antibody levels similar to hospitalized controls and 4.2% (95% CI: 1.4% - 7.4%)) for cases with a distribution of antibody levels similar to outpatient controls. As a result, the fraction of cases in the serosurvey that can be explained with the distribution of the IgG OD ratios from the hospitalized controls, which we refer to as the *indirect indicator of disease severity*, is 51.2% (95% CI: 9.9% − 83.7%). The large 95% CI of this indicator of disease severity is caused by the overlap in the two positive control distributions.

To investigate if the model improves by including a separate estimate for both types of positive controls, we compared the likelihood of the estimates above to the likelihood from a model that is based on either the hospitalized or outpatient control data only (see Equation 1 and Table 1). The p-values in Table 1 indicate that the model is indeed significantly improved by estimating two cumulative incidences separately. Table 1 also shows that the point estimate of the total cumulative incidence estimate is higher if the mixture model is based on the outpatient controls only and lower if it is based on the hospitalized controls only, compared to the model that uses both distributions. This is expected, as the distribution obtained from the COVID-19 hospitalized controls is more distinguishable from the pre-pandemic controls than the COVID-19 outpatient controls.

**Table 1:** Overview of cumulative incidence estimates based on various positive control data. The p-values are the result of a likelihood ratio test.

| Type of positive control data | Cumulative incidence estimate | p-value compared with separate outpatient and hospitalized data |
| --- | --- | --- |
| Outpatient data only | 8.4% (6.9% - 10.1%) | 4.1e-08 |
| Hospitalized data only | 7.7% (6.3% - 9.6%) | 5.0e-07 |
| Outpatient and hospitalized data treated as separate distributions | 8.1% (6.8% - 9.9%) | — |

### Indirect indicator of disease severity differs between age groups

It is known that there is a correlation between the age of an infected individual and the severity of a SARS-CoV-2 infection [21]. To validate our methodology, we estimated the indirect indicator of disease severity for three age-classes: 5 to 40 years, 41 to 65 years and 66 to 90 years. These estimates, together with the total cumulative incidence estimates for the age-classes, are shown in Table 2. Indeed, the indirect indicator of disease severity is highest for the oldest age class: we estimated that 100 % of the cases in the serosurvey can be explained by the distribution of the hospitalized COVID-19 controls, for the middle and young class this is 60.2 % and 21.4 % respectively (see Figure 2). Figure 3 shows that the maximal observed IgG ratio as well as the median of all values above the cutoff provided by the manufacturer (red dots) increase with age. However, the overall median of the distribution does not increase with age (black dots). This illustrates that the observed increase in the indirect indicator of disease severity is driven by the upper part of the IgG ratio distributions. The model that separately considers the age classes is significantly better than the model without these age classes after correcting for the increased amount of parameters (likelihood-ratio test, p-value = 0.009).

**Table 2:** Cumulative incidence and indicator of disease severity for three age-classes.

| Sub-population | Number of observations | Total cumulative incidence | indirect indicator of disease severity |
| --- | --- | --- | --- |
| age-range [5-40] | 1077 | 10.1% (7.8% - 12.7%) | 21.4% (0%-59.6%) |
| age-range [41-65] | 1355 | 7.6% (5.8% - 9.6%) | 60.2% (21.5% - 100%) |
| age-range [66-90] | 334 | 4.1% (0% - 6.9%) | 100% (20.1% - 100%) |
| Females | 1454 | 7.1% (5.3% - 9.0%) | 45.3% (4.1% - 91.9%) |
| Males | 1312 | 9.3% (7.3% - 11.6%) | 50.8% (16.7% - 90.7%) |

**Figure 2:**
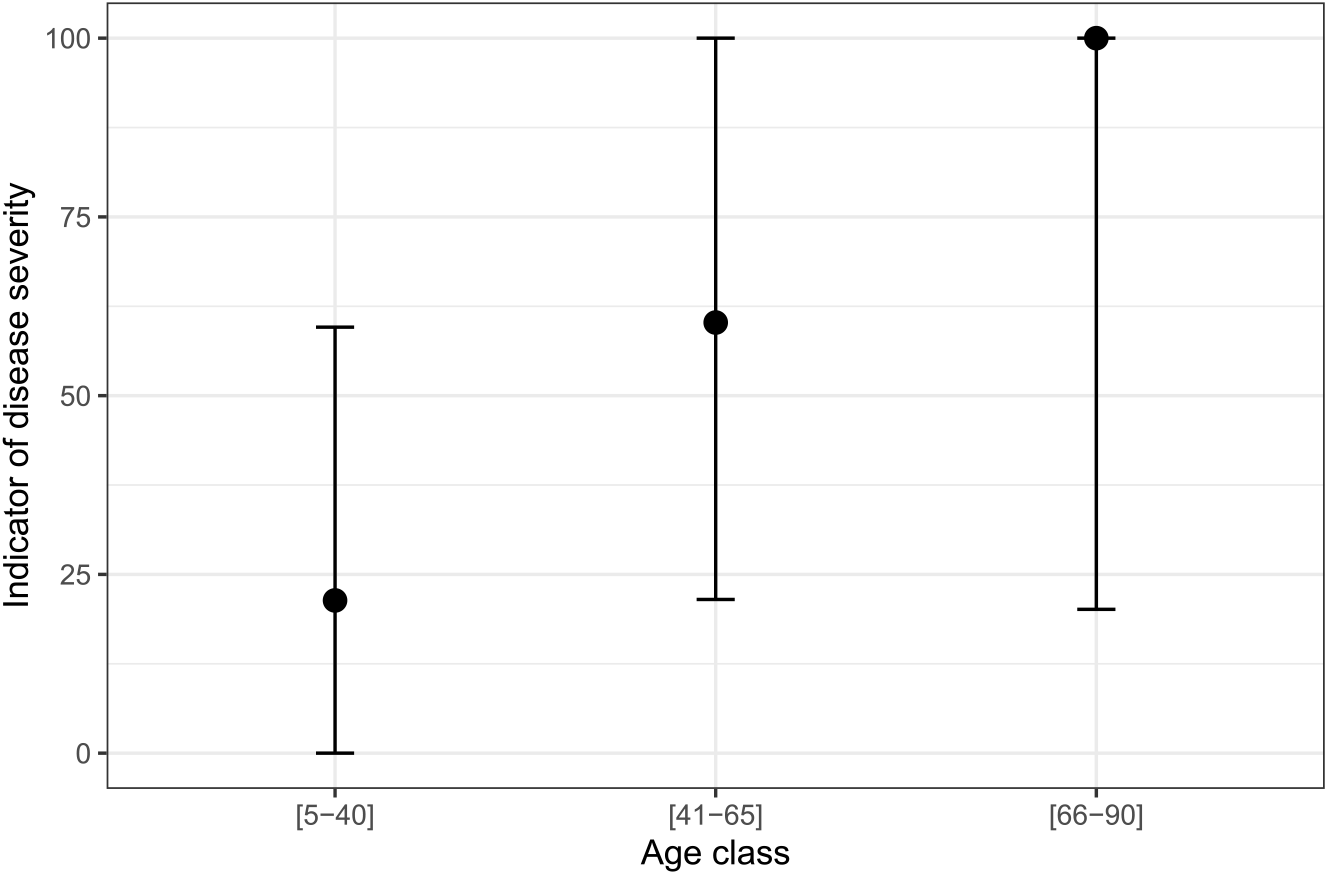
The indirect indicator of disease severity per age class, including the 95% confidence intervals.

**Figure 3:**
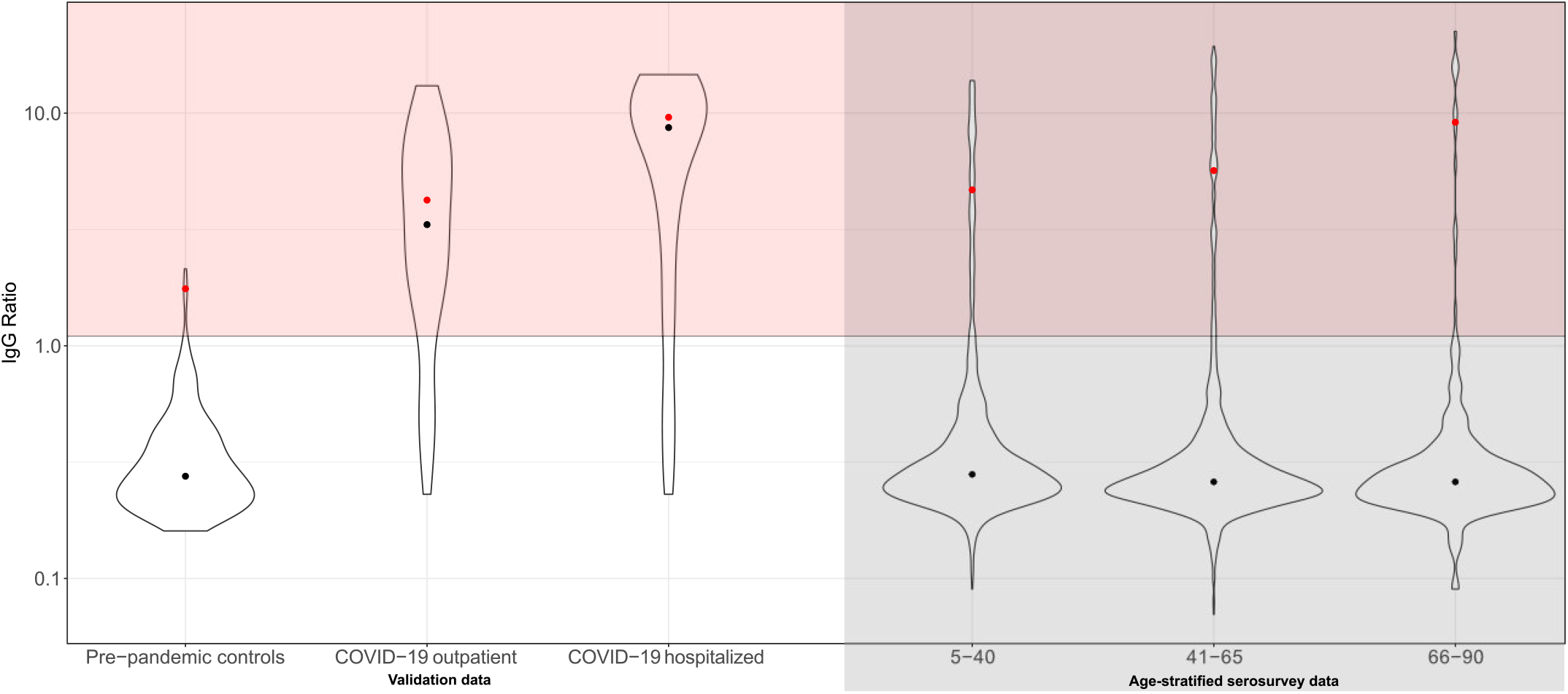
Violin plots of the distributions from the validation data (pre-pandemic controls, COVID-19 outpatient cases and COVID-19 hospitalized cases) and age-stratified serosurvey data. The black dots indicate the median of the full distribution and the red dots the median of all values larger than the cutoff of seropositivity provided by the manufacturer (1.1).

Men, compared to women, are more likely to suffer from a severe SARS-CoV-2 infection [22]. Again, this can also be found by applying the mixture model method to the serosurvey data (see table 2). The point estimate of the indirect indicator of disease severity is higher for males compared to females, although this difference is not significant. The p-value of a likelihood-ratio test for the model that separates female and male participants with the original model is 0.046. The age distribution of the males and females are comparable in the serosurvey (two-sample Wilcoxon test, p-value = 0.18).

### Mismatch between pre-pandemic controls and individuals without previous SARS-CoV-2 infection in the serosurvey

Mixture model methods give unbiased results when the pre-pandemic control data represent individuals without previous COVID-19 infection and the COVID-19 control data span the whole range of COVID-19 severity and their relative occurrence. The method presented in this manuscript can be used to test whether there is a mismatch between the validation and serosurvey data. This is done by testing if there is more statistical support for a extended mixture model that assumes an additional, hidden distribution of antibody levels (cite Bouman pcb and Methods).

In the SEROCoV-POP serosurvey, we indeed infer such a mismatch between the validation and the serosurvey data (p-value likelihood ratio test = 8*e* − 105). Figure 1 shows the distribution of serological measurements that are inferred to be present in the serosurvey data but not in the validation data (purple line – ’additional distribution’). This distribution is on the lower end of the range of antibody levels, covering part of the distribution of the pre-pandemic control samples. This indicates that the inferred mismatch is due to a discrepancy between the measurements from the pre-pandemic control samples and the individuals from the serosurvey study who likely did not have a past SARS-CoV-2 infection.

The total cumulative incidence of SARS-CoV-2 infections for the model that includes the additional case distribution is 9.3% (95% CI: 6.7% - 10.6 %) and thus higher than without this distribution. The reason for this is that the additional distribution is on the lower range of the observed IgG OD Ratio, hence some of the lower range values from the serosurvey data are now inferred to be similar to the additional distribution and thus also COVID-19 cases.

### No evidence for an additional missing positive control distribution with lower mean

The previous subsection describes the mismatch we identified between the negative controls and individuals with low serological measurements in the serosurvey. We hypothesized that there is a further mismatch between the distributions of serological measurements of COVID-19 cases and the serosurvey because the COVID-19 cases in the validation data were all symptomatic and relatively severe and the cases in the serosurvey span the whole spectrum of severity. Specifically, we expected to find evidence for an additional distribution representing COVID-19 cases with a lower mean than the distributions for the outpatient and hospitalized cases.

To test for such an additional case distribution, we lumped all IgG OD ratios below 0.34 into a single point mass to direct the focus of the analysis away from low serological measurements and investigated a potential additional mismatch on the higher end of the observed IgG OD ratios. However, we did not find any statistical support for such an additional mismatch. This suggests that the individuals with high IgG OD ratios in the serosurvey are well represented by the positive control data.

## Discussion

In this study, we present an application of mixture model methods to SARS-CoV-2 serosurvey data. Serosurvey data are currently used to determine the proportion of seropositivity and to estimate the cumulative incidence and the relative risk of seropositivity in various sub-groups. This is usually done by introducing a cutoff for seropositivity.

We show that mixture models that use the entire distribution of the antibody levels rather than a cut-off for seropositivity, provide additional insights into aspects of an epidemic that are usually not addressed in serosurveys. Specifically, we have used mixture models to infer the cumulative incidence from distinct serological distributions, in this case those from hospitalized and outpatient COVID-19 positive controls. We found that the indirect indicator of disease severity (the fraction of individuals with antibody distributions similar to hospitalized cases) increases with age mirroring evidence from clinical studies. Additionally, mixture model methods can be used to test for a mismatch between the pre-pandemic and COVID-19 control data and the serosurvey data, which could indicate that the cases observed in the population are not well represented by those included in the control data. While we provide evidence for such discrepancies, they are not indicative of a large fraction of cases with intermediate antibody levels that would be expected for asymptomatically infected individuals.

Other studies using mixture model methods to analyse SARS-CoV-2 serosurvey data have been conducted. Vos et al (2021) used mixture models to validate their cutoff value [23]. They assumed and inferred control and case distributions from the serosurvey data [23]. Our approach in contrast, is based on the observed distributions of serological measurements in prepandemic sera and sera of individuals with PCR-confirmed SARS-CoV-2 infection. These observed distributions are not adequately captured by the normal distributions Vos et al assumed. Hence, our study represents a more stringent and empirically-supported use of mixture models.

Although the mixture model approach naturally allows to implement declining antibody levels and seroreversion [24], we have not corrected our estimate of the cumulative incidence for the possible effect of sero-reversion. The reason for this is that the serosurvey was conducted within 4 months of the start of the pandemic. Current estimates of antibody half lives IgG RBD are around 50–106 days [25]. Therefore we expect the effect of sero-reversion to be negligible. Furthermore, we did not correct the estimate of cumulative incidence for age nor household structure because our study was aiming to provide a proof of concept rather than additional estimates for the sero-prevalence in Geneva. As a result, the estimates presented here are only representative for the study population and not for the general population of Geneva. Estimates for the cumulative incidence of the general population of Geneva from these data can be found in Stringhini et al. (2020) [2].

The presented estimates of the indirect indicator of disease severity have wide confidence intervals. This is caused by the fact that while the distributions of the antibody levels for COVID-19 hospitalized and outpatient cases are significantly different, there is quite a lot of overlap (see figure S1). This could potentially be improved if more detailed positive control data would be available to guide the construction of more distinguishable distributions of IgG OD ratios based on characteristics of the infections or infected individuals. Despite the large confidence intervals, we found that the indirect indicator of disease severity increases with age, corroborating previous reports [21]. Similarly, the point estimate of the indirect indicator of disease severity is higher for males compared to females, consistent with reported sex differences in ICU admission and death [22].

Mixture model methods give unbiased results when both the negative and positive controls in the validation data represent the general population well [8]. However, we know that there are some issues with our validation data. First, the pre-pandemic control data over-represent individuals with pathological conditions and, for both the pre-pandemic and the COVID-19 controls, the age-distribution is different from the general population [14]. Second, all cases in the COVID-19 control group show at least mild symptoms and half of them have been hospitalized. Therefore, the severity of the selected cases is higher than expected in a random group of COVID-19 patients and is not consistent with the estimated fraction of 20 % asymptomatic cases [26]. This is likely to result in a different distribution of serological measurements as severe cases have been shown to give rise to higher antibody levels than mild cases [19], [27]. Third, the ratio of outpatient to hospitalized cases in the control group is lower than expected, which could lead to an underestimation of the cumulative incidence [8]. More extensive and representative validation data could improve the cumulative incidence estimates, however, such data are difficult to collect, especially at the beginning of a pandemic, when asymptomatic and mild cases often go undetected and their proportion is unknown.

Part of the aim of our approach was to identify potential biases caused by any of the mentioned limitations in the validation data. Interestingly, however, the mismatch we identify is not characterized by an intermediate level of antibodies in between the level of the pre-pandemic sera and the outpatients as we would expect for a missing distribution of mild or asymptotic cases. Opposite to our expectation we found that the serosurvey data display a narrower distribution at the lower end of the antibody levels than the pre-pandemic, negative controls — as if there were asymptotic or mild SARS-CoV-2 infections among the pre-pandemic controls. A more detailed characterization of the individuals from whom the pre-pandemic control sera were sampled, as well as the determination of antibody levels in asymptomatic and mild cases could shed further light on this mismatch and thus further improve the estimation of the cumulative incidence. An additional improvement could be obtained when a quantitative immuno-assay would be used, instead of the semi-quantitative Euroimmun that was available at the beginning of the pandemic.

## Data Availability

Data are available upon reasonable request.

## Acknowledgements

We would like to thank Peter Ashcroft, Sonja Lehtinen and Jana Huisman for valuable comments on the manuscript. Roland Regoes gratefully acknowledges funding from the Botnar Research Centre for Child Health (grant number 2020-FS-354).

## Ethics Statement

The SEROCoV-POP study was approved by the Cantonal Research Ethics Commission of Geneva, Switzerland (CER16-363). The full study protocol is available online (in French).

## Supplementary Material

**Figure S1:**
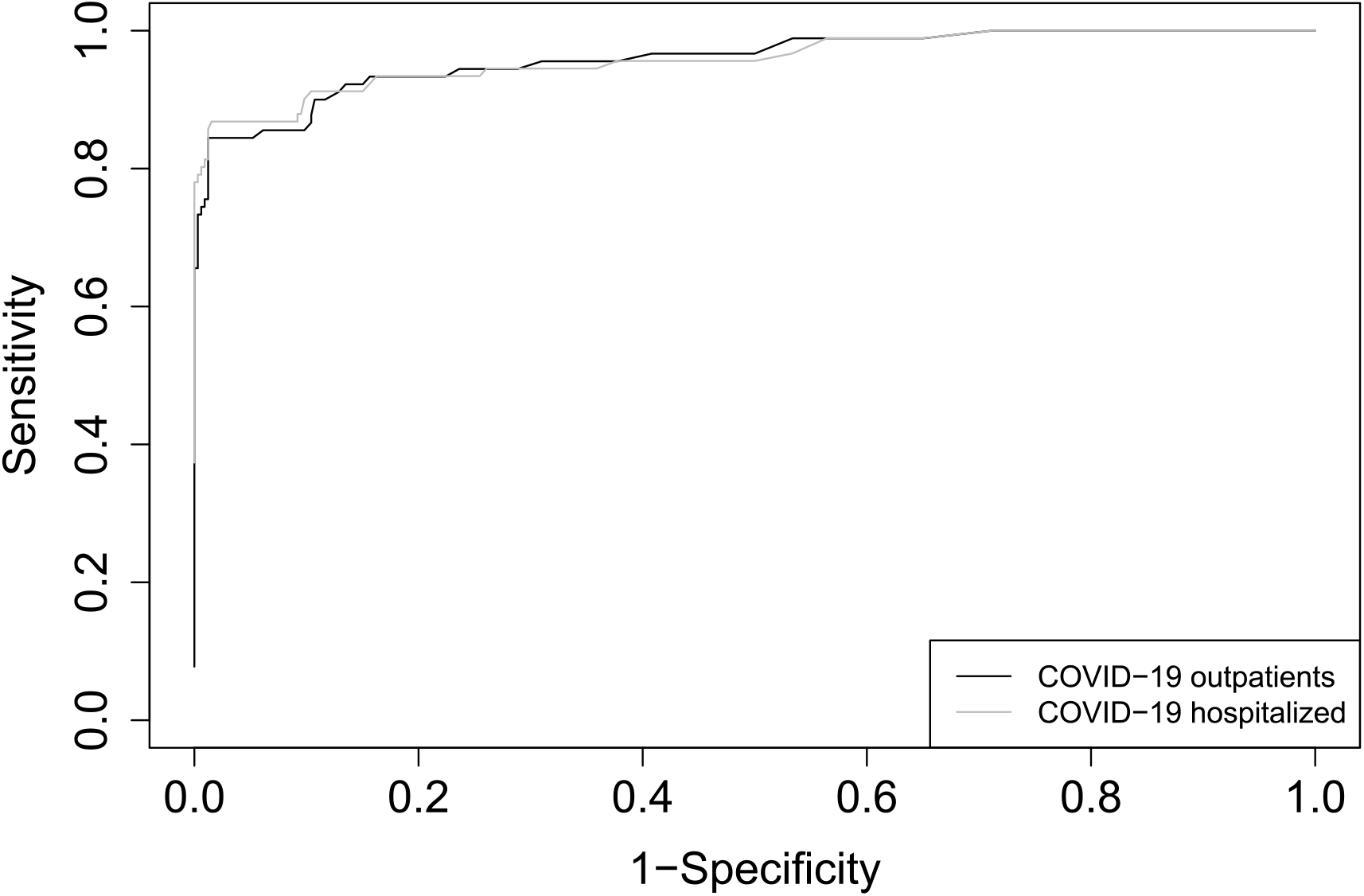
ROC-curve of the validation data for both the COVID-19 outpatient controls (black) and COVID-19 hospitalized controls (grey).

